# Predicting Positive Psychological States using Machine Learning and Digital Biomarkers from Everyday Wearable Data

**DOI:** 10.1101/2025.04.26.25326492

**Authors:** Lingxi Wu, Arlene John, Jorge Piano Simões

## Abstract

Wearable devices offer continuous physiological data collection, presenting new opportunities for real-world mental health monitoring. Previous research has primarily emphasized detecting stress and psychological states associated with mental illnesses, whereas predicting positive psychological states, such as self-esteem, positive affect, and meaning in life, remains underexplored.

In this study, 34 participants wore research-grade smartwatches over an eight-day period, resulting in 6,528 hours of physiological data—including heart rate variability (HRV), electrodermal activity (EDA), and accelerometer-derived movement features. Psychological states were concurrently assessed using Ecological Momentary Assessment (EMA), yielding 247 observations with a total of 4,446 self-reported labels across 18 psychological states related to positive and negative affect, self-esteem, sense of meaning in life, and personal relationships. Machine learning models—including Random Forest, Long Short-Term Memory networks (LSTM), Convolutional Neural Networks (CNNs), and Transformers—were trained to predict these EMA-reported psychological states. Explainable AI techniques (SHAP and LIME) identified physiological markers of importance, while conformal prediction methods assessed model uncertainty.

Results indicate that absolute prediction accuracy remains challenging; however, CNN models achieved threshold accuracies of up to 62%, with accelerometer-based features emerging as most predictive. Self-esteem-related states demonstrated the clearest physiological signatures, and among negative affect states, anger was more reliably detectable than anxiety or sadness.

These findings highlight the potential of wearable-derived biomarkers for monitoring positive psychological states, despite current limitations in predictive accuracy. Future research should focus on refining feature extraction methods, enhancing model generalizability, and integrating multimodal data sources to improve real-world applicability in mental health monitoring.

## Introduction

In recent years, wearable devices have significantly advanced the collection and analysis of physiological and behavioral data in psychological research. Smart devices like smartwatches, trackers, and rings can continuously track physiological metrics like heart rate, skin conductance, physical activity, and sleep patterns [1]. These continuous measures facilitate unprecedented insights into health and psychological states within naturalistic, real-world contexts [2]. This technological progression has driven the emergence of digital phenotyping, defined as the moment-by-moment quantification of individual-level human behavior and experiences through mobile devices and wearable technology [3]. This integration of objective, unobtrusive physiological data via digital phenotyping offer transformative potential for psychological research, enabling objective measurement of subjective states such as emotional experiences, personality characteristics, and cognitive capacities [4].

A particularly impactful application of digital phenotyping combined with machine learning has been in the detection and monitoring of mental health disorders. Indeed, recent systematic literature reviews highlight a growing interest in using these technologies to detect conditions such as stress, anxiety, and depression, which represent major global health priorities due to their significant personal, social, and economic impact [1, 5]. However, the applications of digital phenotyping need not be limited to pathological states; these methodologies also offer promising opportunities for understanding broader aspects of mental health.

For instance, digital phenotyping and machine learning approaches can capture fluctuations in everyday psychological states that are present irrespective of clinical diagnosis. Ecological momentary assessment [6], which involves repeated sampling of individuals’ subjective states in their natural environment, is particularly valuable in this context. EMA enhances ecological validity by collecting data in real-world settings, provides rich contextual information about the circumstances surrounding psychological experiences, and allows longitudinal tracking of within-person fluctuations over time [7]. This approach facilitates capturing variations in positive psychological states, such as self-esteem, personal meaning, quality of interpersonal relationships, and the balance of positive versus negative affect, all of which have been shown to operate as state-like phenomena in previous EMA research [8]. These dimensions, central to positive psychology and well-being research, intersect with clinical perspectives through frameworks such as the dual-continuum model [9], which conceptualizes mental health and mental illness as related yet distinct phenomena. Thus, extending digital phenotyping beyond symptomatology towards the assessment of positive mental health states can expand the current understanding of mental health as a continuum, offering complementary insights to both clinically and in everyday contexts.

Although machine learning methods have enabled powerful predictive models in psychological research, concerns regarding their interpretability and transparency have historically limited their broader integration into routine care or daily life. Recent advancements in explainable artificial intelligence (XAI) now provide tools to quantify and interpret the inner workings of complex models. Techniques such as feature importance analysis, SHAP values, and prediction intervals can elucidate the factors driving model predictions, thus increasing transparency, trustworthiness, and ultimately facilitating their practical adoption in clinical and everyday settings [10].

To explore the utility of digital phenotyping for capturing positive mental health states, we conducted an observational study integrating wearable smart devices and EMA. Our study specifically focused on capturing fluctuations in key positive psychological constructs, including self-esteem, positive affect, personal meaning, and the quality of interpersonal relationships. By combining continuous physiological monitoring and context-rich subjective assessments collected via EMA, we aimed to establish robust, ecologically valid methods for assessing everyday positive mental health, thus extending the traditional boundaries of psychological research and practice.

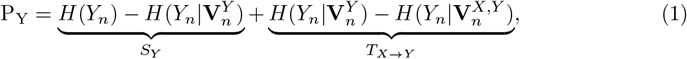

## Materials and methods

### 1 Dataset

#### 1.1 Dataset Overview

The study was conducted under the approval of the University of Twente Ethics Committee. The Ethics Committee of the University of Twente’s Faculty of Behavioural Sciences approved this study (project 240133, obtained on 23/02/2024). This dataset offers a rich combination of physiological and psychological data, uniquely suited for studying the relationship between wearable signals and mental states in young adults. While the dataset is relatively small compared to those used by large technology companies, it provides a controlled and ecologically valid foundation for academic research.

#### 1.2 Participant Demographics

The dataset was collected by the PHT (Psychology, Health, and Technology) department in the BMS (Behavioural Management and Social Sciences) faculty at the University of Twente. Data collection occurred between March 19, 2024, and May 13, 2024, involving 34 participants. The participants were university students aged 18–24, with a gender distribution of 37% men and 63% women. Recruitment was conducted via university-wide announcements, and all participants provided written informed consent prior to their involvement in the study.

#### 1.3 Data Collection Procedure

Participants were equipped with Empatica Embrace Plus wearable devices[11], which continuously monitored their physiological signals and activity patterns throughout the study period of 8 days. The devices recorded multiple modalities, including: **HRV:** Capturing cardiovascular activity, **EDA:** Measuring skin conductance as an indicator of emotional arousal, **ACC:** Detecting movement and activity levels. Participants were instructed to wear the smartwatch on their non-dominant hand to minimize interference with daily activities while ensuring consistent data collection. However, individual differences in variations in movement patterns could introduce inconsistencies in accelerometer (ACC) measurements. Given that ACC-derived features are highly sensitive to motion artifacts, variations in wrist positioning may affect feature accuracy, particularly for activity intensity and entropy-based movement metrics. Participants were instructed to answer a questionnaire at the end of the day. The questionnaire is connected to the concepts of P:positive effects, N: negative effects, SE: self-esteem, MIL: meaning in life, and PRO: Personal Relationships, and is shown in Table 1.

**Table 1.** Survey Questions with Codes and LabelS.

| Label | Code | Question |
| --- | --- | --- |
| 0 | P1 | I felt joyful today. |
| 1 | N1 | I felt anxious today. |
| 2 | P2 | I felt positive today. |
| 3 | N2 | I felt angry today. |
| 4 | N3 | I felt sad today. |
| 5 | P3 | I felt content today. |
| 6 | SE1 | On the whole, I was satisfied with myself today. |
| 7 | SE2 | I was inclined to feel like a failure today. |
| 8 | SE3 | I certainly felt useless today. |
| 9 | SE4 | I feel that I had a number of good qualities today. |
| 10 | MIL1 | Today I understood my life's meaning. |
| 11 | MIL2 | Today I was searching for meaning in life. |
| 12 | MIL3 | Today my life was full of value. |
| 13 | MIL4 | Today I was highly committed to certain goals in my life. |
| 14 | MIL5 | Today I could comprehend what my life is all about. |
| 15 | PRO1 | Today I received help and support from others when I needed it. |
| 16 | PRO2 | Today I felt loved. |
| 17 | PRO3 | Today I was satisfied with my personal relationships. |

### 2 Dataset processing

To transform the raw wearable data into machine-learning-ready features, a structured pipeline was implemented, as illustrated in Figure 1. The steps involved in this process are described below.

**Fig 1.**
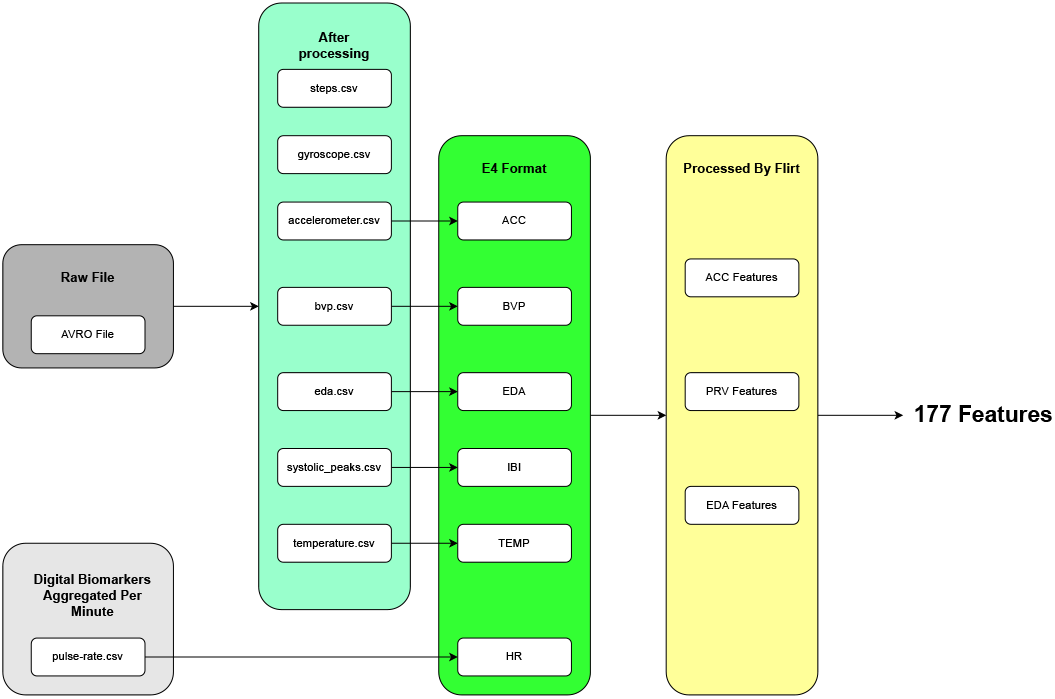
Workflow for Data Preprocessing and Feature Engineering. Raw data was processed, transformed to E4 format, and feature-engineered using the FLIRT toolkit.

#### 2.1 Data preprocessing

This section describes the signal processing workflow implemented in Python for extracting physiological data from .avro files. The data collected by the embraceplus can be downloaded from the cloud as .avro files [12]. The data consists of multiple physiological modalities such as ACC, gyroscope, EDA, temperature, and more. To illustrate the nature of these signals, figure 2 shows a 1-day segment of raw data for each modality. The *Empatica Documentation* [13] was consulted to ensure the data adhered to the expected standards and formats for physiological signal processing. First, the raw digital values into meaningful physical units, and precise timestamps based on the sampling frequency and start time were calculated. The processed data is then saved in a structured format for subsequent analysis. Finally, individual sensor streams were extracted from the AVRO files and saved as regular files, including Step counts, Gyroscopic motion data, ACC data, Blood volume pulse data, EDA data, detected systolic peaks for calculating inter-beat intervals (IBI), and skin temperature. The inter-beat intervals (IBI) were computed by taking the difference between consecutive systolic peaks. Pulse filtering was optionally applied for further refinement and pulse-rate aggregated digital biomarkers (e.g., heart rate) per minute.

**Fig 2.**
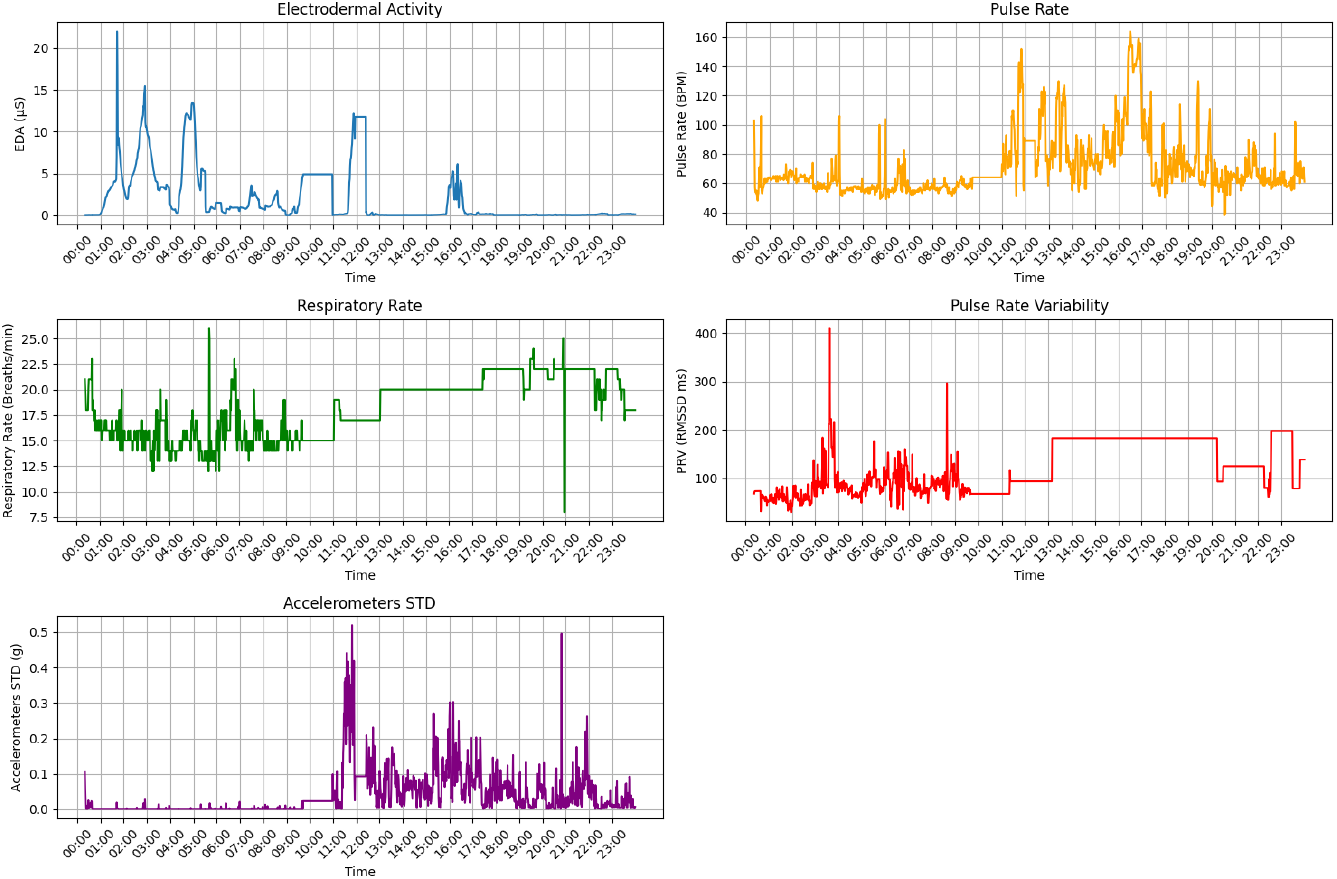
Example raw data for each modality

#### 2.2 Adjustments for FLIRT Toolkit Compatibility

FLIRT is a feature generation toolkit designed to extract interpretable, high-quality features from multimodal wearable sensor data using time-, frequency-, and statistical-domain transformations [14]. It streamlines the preprocessing and feature engineering pipeline for time-series health and activity recognition tasks, enabling reproducible and scalable machine learning workflows. The FLIRT toolkit is optimized for Empatica E4 data formats, which is the previous generation of Empatica product family and does not natively support the latest Empatica Embrace Plus model. Therefore, adjustments were necessary to ensure the processed files adhered to the FLIRT-compatible input format.

The modifications ensure that the processed data can match the FLIRT input requirements, also retain data fidelity during transformations and allow for seamless feature extraction using FLIRT. FLIRT was utilized to extract a comprehensive set of features from the processed data. FLIRT uses an overlapping sliding-window approach to generate machine-learning-ready features. Key feature categories include:

##### ACC Features

Statistical and time-series features derived from accelerometer data, capturing movement intensity and patterns.

##### PRV Features

Features derived from IBI data, providing metrics for pulse rate variability, which serves as a proxy for heart rate variability.

##### EDA Features

Statistical features calculated from electrodermal activity, which reflect physiological responses such as stress and arousal.

The supplementary materials includes a list of the features generated by FLIRT based on various physiological signals. In total, 178 features are extracted, aggregated over a one-hour timestep and time window.

#### 2.3 Dataset preparation

The generated features were further processed to ensure consistency and readiness for further analysis. The preprocessing steps included the removal of corrupted entries, interpolation of missing values, and standardization where necessary.

The preprocessing steps carried out included the removal of columns/features with excessive missing values and imputing missing values for the remaining columns using the median. Data from all participants were compiled into a unified dataset and residual missing and infinite values for checked to ensure data integrity.

##### 2.4 Scaling and PCA

After preprocessing the raw dataset, the data was split into three versions for analysis:

##### Original Dataset

Retained the original features without further transformations.

##### Scaled Dataset

Standardized the features to have zero mean and unit variance:H

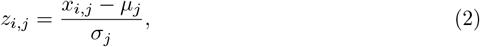

where *x*_*i,j*_ is the original value, *µ*_*j*_ is the mean, and *σ*_*j*_ is the standard deviation of the *j*-th feature.

##### PCA Dataset

Applied Principal Component Analysis (PCA) to the already scaled dataset to reduce dimensionality, retaining 95% of the explained variance:

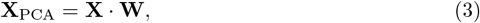

where **W** is the matrix of principal components.

The dataset was divided into training and testing subsets for each label. To ensure no data leakage between participants, a grouped split was applied based on participant IDs. This guarantees that data from the same individual does not appear in both training and testing sets

#### 2.5 Feature selection analysis

The extracted features were examined for relationships and potential redundancies. t-SNE, correlation analysis and feature importance were assessed to understand the interrelationships among the features, ensuring the dataset’s quality for downstream machine learning tasks.

#### 2.6 Machine learning Models

This section describes the experimental methodology for training and evaluating machine learning models to predict each label in the dataset. The experiments were conducted in three stages: dataset preparation, model training, and result comparison.

##### Models and Training Procedure

Various kinds of machine learning models were trained separately in the dataset:

##### Random Forest (RF)

A tree-based ensemble model used for classification tasks [15].

##### Convolutional Neural Network (CNN)

A deep learning model designed to learn spatial hierarchies in the feature space:

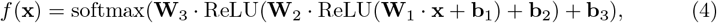

where **W**_*i*_ and **b**_*i*_ are the weights and biases of the *i*-th layer [16].

##### Long Short-Term Memory (LSTM)

A recurrent neural network capable of capturing temporal dependencies in sequential data:

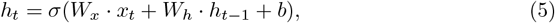

where *h*_*t*_ is the hidden state at time *t*, and *σ* is the activation function [17].

##### XGBoost (XGB)

A gradient boosting framework designed for high-performance tree-based learning tasks:

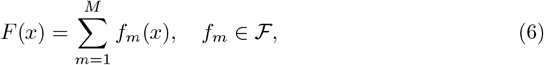

where each *f*_*m*_(*x*) is a regression tree and ℱ is the space of all possible trees [18].

##### LightGBM (LGBM)

A gradient boosting algorithm optimized for efficiency and scalability using a histogram-based learning approach:

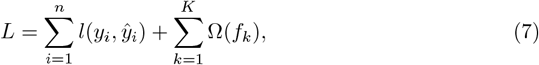

where *l*(*y*_*i*_, *ŷ*_*i*_) is the loss function, and Ω(*f*_*k*_) represents the regularization term for tree complexity [19].

#### 2.7 Modelling Protocol

After data cleaning, standardization was applied to normalize the feature values across the dataset. Principal Component Analysis (PCA) was then performed to reduce dimensionality while retaining the most informative components. To ensure the reliability and robustness of the models, a 5-fold cross-validation strategy was employed.

Subsequently, five machine learning models—1D Convolutional Neural Network (1D-CNN), 1D Long Short-Term Memory (1D-LSTM), LightGBM (LGBM), Random Forest, and XGBoost—were implemented and evaluated across the original dataset, the scaled dataset, and the PCA feature-dataset. The input data consisted of one-dimensional arrays representing all extracted features within an hourly window. The task was formulated as a classification problem, where each response was categorized into one of 11 classes, corresponding to integer values ranging from 0 to 10. Each of the 18 survey questions was treated as a separate prediction target, with models trained and evaluated independently for each label. To align the question responses with the extracted hourly features, answers from the same day were assigned to the final timestamp within the corresponding sequence. However, due to the nature of the problem, treating each one-dimensional feature array within an hourly window as an independent input to predict the same daily label led to the loss of crucial temporal information. To address this issue, the features were subsequently transformed into two-dimensional arrays, preserving their original temporal structure. Padding was applied to ensure uniform sequence lengths across all samples.

#### 2.8 Metrics

For each model, the training set was used to fit the model, while the testing set evaluated performance. Metrics such as accuracy, top-3 accuracy, and threshold accuracy were calculated as follows:

##### Accuracy

Accuracy is a common evaluation metric for multi-class classification tasks that measures the proportion of correctly predicted instances out of the total number of instances.

Let *N* be the total number of instances, *y*_*i*_ be the true class label for the *i*-th instance, *ŷ*_*i*_ be the predicted class label for the *i*-th instance. Then, the accuracy is defined as:

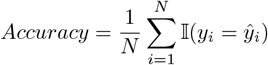

where 𝟙(*·*) is the indicator function, which returns 1 if the argument is true and 0 otherwise.

##### Top-3 Accuracy

The *Top-3 Accuracy* measures the proportion of predictions where the true label *y*_*i*_ is among the top three labels {*ŷ*_*i*,1_, *ŷ*_*i*,2_, *ŷ*_*i*,3_} predicted by the model. It is defined as:

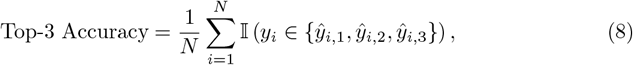

where *N* is the total number of samples. *ŷ*_*i*,1_, *ŷ*_*i*,2_, *ŷ*_*i*,3_ are the top three predicted labels for sample *i*, ranked by confidence and 𝟙 () is an indicator function that equals 1 if the condition is true and 0 otherwise.

This metric reflects the model’s ability to make correct predictions within its top three guesses, which is particularly useful when the labels represent approximate states or when slight variations in ranking are acceptable.

##### Threshold Accuracy

The *Threshold Accuracy* evaluates the proportion of predictions *ŷ*_*i*_ that fall within a specific tolerance range around the true label *y*_*i*_. For a label range of [0, 10], with a threshold tolerance of *±*1, it is defined as:

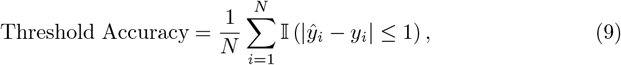

where |*ŷ*_*i*_ −*y*_*i*_| ≤1 ensures the prediction *ŷ*_*i*_ is within one unit of the true label *y*_*i*_. All other terms are as previously defined. This metric is meaningful in scenarios where predictions close to the true label are as valuable as exact matches. For example, predicting a “stress level” of 5 instead of the true value of 6 might still provide actionable insights.

Each model’s weights were saved for reproducibility. By evaluating both *Top-3 Accuracy* and *Threshold Accuracy*, we assess the model’s ability to make accurate ranked predictions (*Top-3 Accuracy*) and predict labels within a reasonable tolerance range (*Threshold Accuracy*).

#### 2.9 Interpretability

To enhance the interpretability of the machine learning models, Explainable AI (XAI) techniques were employed. These methods enable a deeper understanding of model behavior and the contributions of individual features:

##### MAPIE (Mean and Prediction Interval Estimation)[20]

MAPIE (Mean and Prediction Interval Estimator) is a model-agnostic tool that enhances explainability by providing both point predictions and calibrated prediction intervals, allowing users to assess not just what the model predicts but how confident it is in those predictions.

This technique was utilized to quantify the uncertainty of predictions by providing confidence intervals, thereby enhancing the reliability of the model’s outputs.

##### SHAP (SHapley Additive exPlanations)[21]

SHAP (SHapley Additive exPlanations) is an explainability method that quantifies the contribution of each feature to a model’s prediction by computing Shapley values from cooperative game theory. SHAP values were computed to assess the contribution of each feature to the model’s predictions. This method provided both global insights into feature importance and local interpretability for individual predictions.

## Results

### 2.10 Data collection results

The dataset includes 272 days of data, amounting to 6528 hours of recordings, with each participant contributing 192 on average. The demographics of the participants in the study are shown in Table 2. In addition to physiological data, the dataset contains metadata such as timestamps, gender, nationality.etc. To ensure privacy and ethical compliance, all data was anonymized, with participant IDs replacing personal identifiers. Figure3 shows the distribution of answers as labels.

**Table 2.** Sociodemographic characteristics of participants.

| Baseline Characteristics | Participant information (n=34) |
| --- | --- |
| Gender (male:female) | 1:1 |
| Age (Mean ± std) | 21.45 ± 2.05 |
| Occupation: |  |
| Student | 17 |
| Studying and Working | 12 |
| Working | 5 |
| Nationality: |  |
| German | 32 |
| Dutch | 1 |
| Vietnamese | 1 |

**Fig 3.**
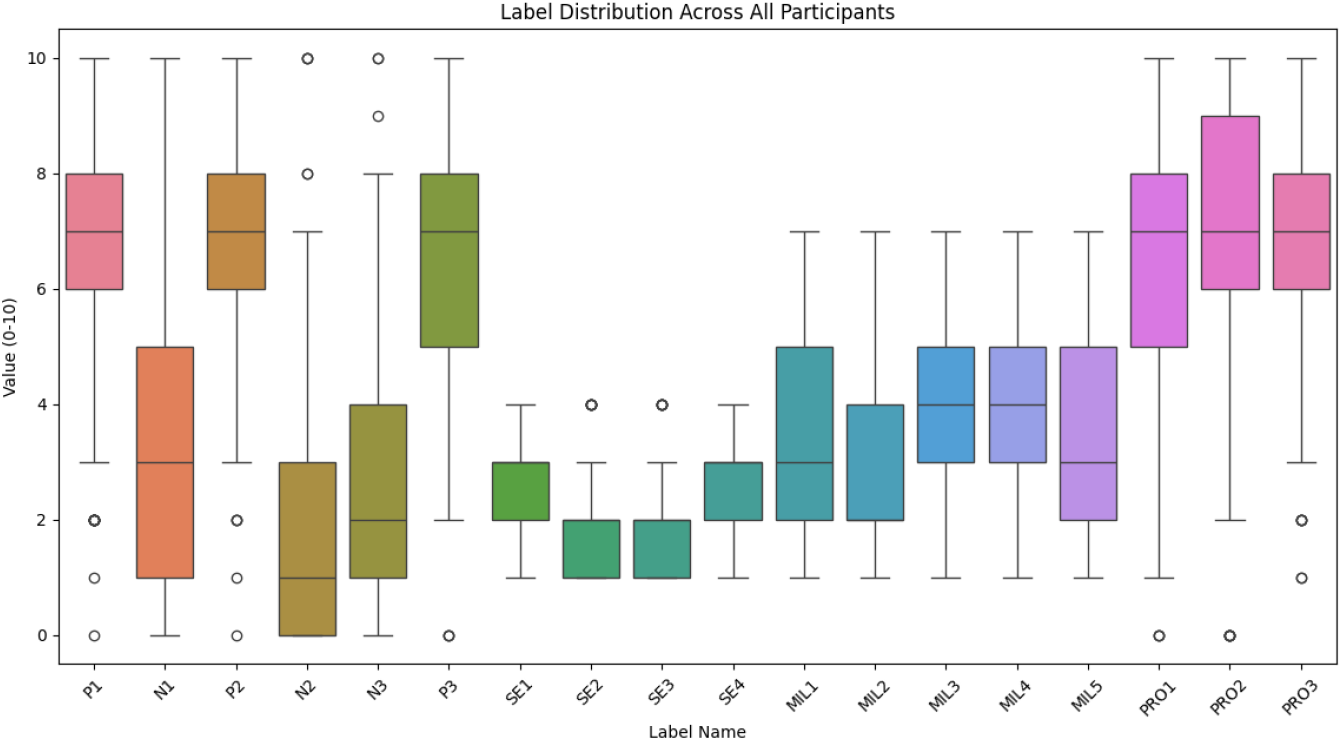
Label Distribution of answers to questions in the survey, range from 0 to 10. P: Positive Affect, N: Negative Affect, SE: Self-Esteem, MIL: Meaning in Life, PRO: Personal Relationships

### 2.11 Feature analysis

#### 2.11.1 t-SNE

Figure 4 presents a t-distributed Stochastic Neighbor Embedding (t-SNE) visualization of the extracted features, where each point represents a feature vector from the dataset. The points are color-coded based on the dominant sensor modality:

**Fig 4.**
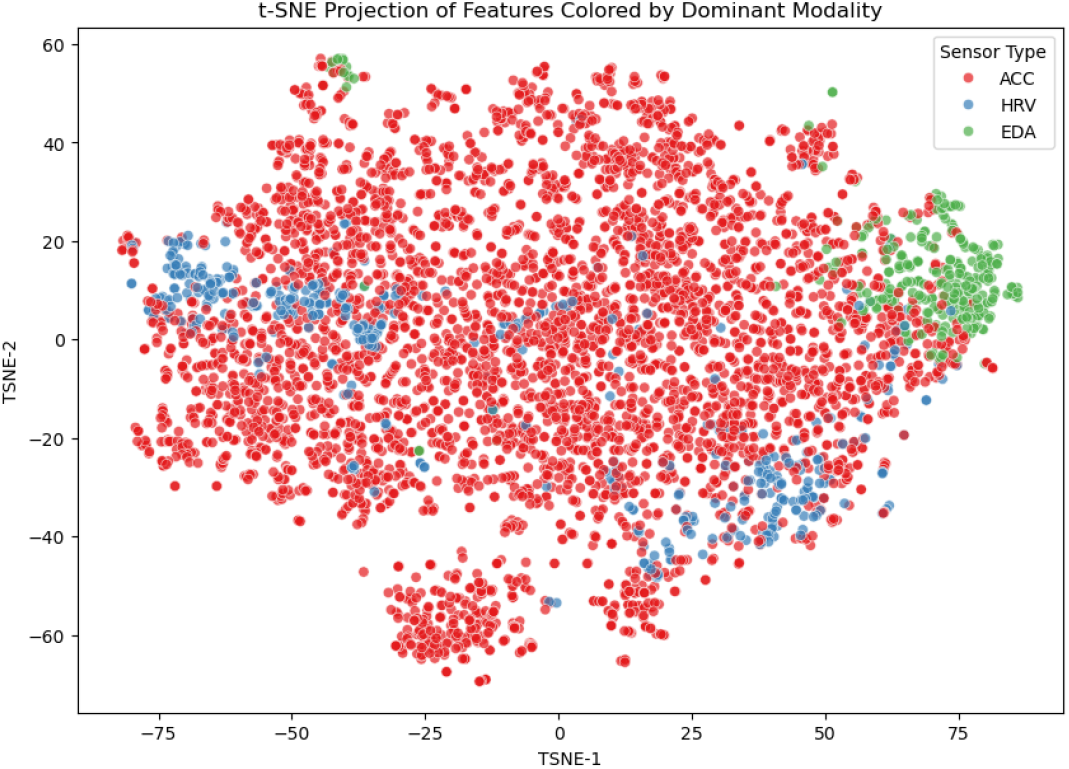
t-SNE Projection of Features Colored by Dominant Modality

- **Red (ACC - Accelerometer)**
- **Blue (HRV - Heart Rate Variability)**
- **Green (EDA - Electrodermal Activity)**

The distribution of feature points provides insights into the clustering behavior of different sensor modalities:

- **ACC features (red)** dominate the feature space, suggesting that the majority of extracted features are derived from accelerometer data.
- **EDA features (green)** form a distinct cluster in the upper-right region, indicating that they capture unique information different from ACC and HRV.
- **HRV features (blue)** are scattered throughout the space, with some overlap with both ACC and EDA, suggesting that HRV-based features share similarities with the other modalities but do not form a strongly independent group.

This visualization helps to understand the relationship between different sensor modalities and how distinct or overlapping their feature representations are.

#### 2.11.2 Feature Correlation

Figure 5 illustrates the mean correlation coefficient between features derived from different sensor modalities. The following modality pairs are analyzed:

**Fig 5.**
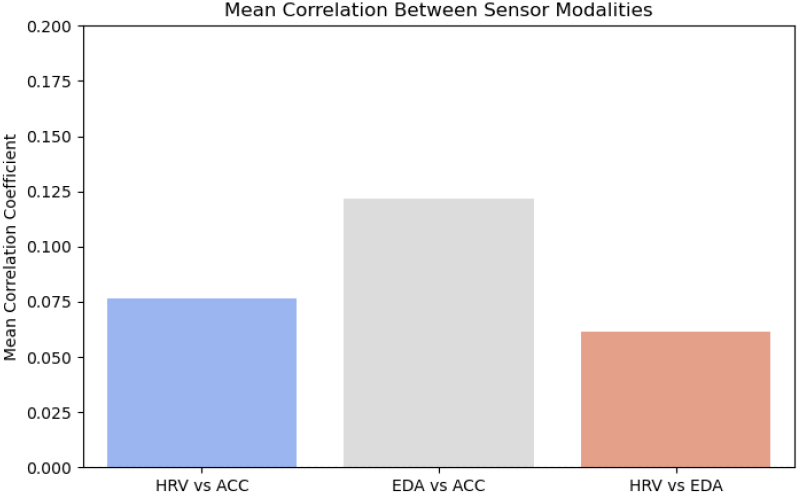
Mean Correlation Between Sensor Modalities

- **HRV vs. ACC (blue bar)**: Displays a relatively low correlation, suggesting that HRV and accelerometer features capture largely independent information.
- **EDA vs. ACC (gray bar)**: Shows the highest correlation, indicating that electrodermal activity and accelerometer data share some common patterns, possibly due to movement-related artifacts in EDA signals.
- **HRV vs. EDA (red bar)**: Exhibits the lowest correlation, confirming that HRV and EDA features are the most independent, as expected given their distinct physiological processes.

This correlation analysis helps determine the level of redundancy or complementarity among sensor modalities. The low correlation between HRV and EDA suggests that combining them in predictive models may provide richer and more diverse information, improving classification or regression tasks. Conversely, the higher correlation between EDA and ACC suggests that these modalities might contribute redundant information if not handled properly.

##### Key Observations

- The t-SNE visualization shows that ACC features dominate the dataset, while EDA forms a distinct cluster, and HRV features are more dispersed.
- The correlation analysis indicates that EDA and ACC features have the strongest relationship, while HRV and EDA features are the most independent.
- These findings suggest that combining features from different sensor modalities, particularly HRV and EDA, may provide complementary information and improve model performance.

#### 2.11.3 Model Performance per label

Table 3 shows the performance comparison between models and datasets.

**Table 3.** Performance Metrics (F1 Score, Threshold Accuracy, Top 3 Accuracy)

| Model | Dataset | Accuracy | Threshold Accuracy | Top 3 Accuracy |
| --- | --- | --- | --- | --- |
| CNN | original | $0.2795 \pm 0.0359$ | $0.6112 \pm 0.0447$ | $0.6443 \pm 0.0465$ |
| CNN | pca | $0.2200 \pm 0.0194$ | $0.5446 \pm 0.0224$ | $0.5776 \pm 0.0245$ |
| CNN | scaled | $0.2169 \pm 0.0221$ | $0.5396 \pm 0.0304$ | $0.5755 \pm 0.0313$ |
| LGBM | original | $0.2305 \pm 0.0259$ | $0.5524 \pm 0.0356$ | $0.6006 \pm 0.0348$ |
| LGBM | pca | $0.2325 \pm 0.0257$ | $0.5529 \pm 0.0347$ | $0.5970 \pm 0.0310$ |
| LGBM | scaled | $0.2304 \pm 0.0250$ | $0.5528 \pm 0.0380$ | $0.5992 \pm 0.0336$ |
| LSTM | original | $0.2598 \pm 0.0334$ | $0.5910 \pm 0.0408$ | $0.6222 \pm 0.0369$ |
| LSTM | pca | $0.2674 \pm 0.0305$ | $0.5909 \pm 0.0384$ | $0.6352 \pm 0.0423$ |
| LSTM | scaled | $0.2643 \pm 0.0321$ | $0.5960 \pm 0.0419$ | $0.6318 \pm 0.0352$ |
| Random Forest | original | $0.2429 \pm 0.0299$ | $0.5753 \pm 0.0404$ | $0.5338 \pm 0.0285$ |
| Random Forest | pca | $0.2566 \pm 0.0286$ | $0.5883 \pm 0.0377$ | $0.5491 \pm 0.0224$ |
| Random Forest | scaled | $0.2441 \pm 0.0310$ | $0.5762 \pm 0.0414$ | $0.5338 \pm 0.0275$ |
| XGB | original | $0.2295 \pm 0.0244$ | $0.5545 \pm 0.0341$ | $0.5934 \pm 0.0303$ |
| XGB | pca | $0.2302 \pm 0.0224$ | $0.5565 \pm 0.0330$ | $0.5932 \pm 0.0345$ |
| XGB | scaled | $0.2299 \pm 0.0246$ | $0.5538 \pm 0.0340$ | $0.5925 \pm 0.0309$ |

Figures 6, 7, and 8 illustrate the performance of the Random Forest, LSTM, and CNN models evaluated on three datasets: the original dataset, the dataset with PCA-applied features, and the scaled dataset. The performance was assessed using three metrics: accuracy, Top-3 accuracy, and threshold accuracy.

**Fig 6.**
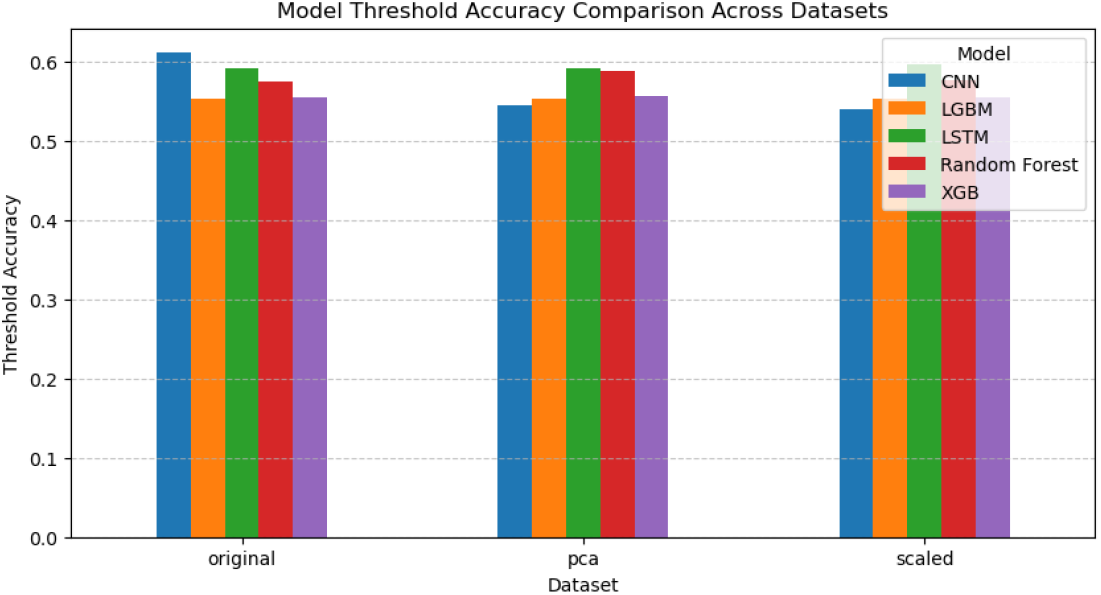
Accuracy Comparison of Random Forest, LSTM and CNN in 3 datasets

**Fig 7.**
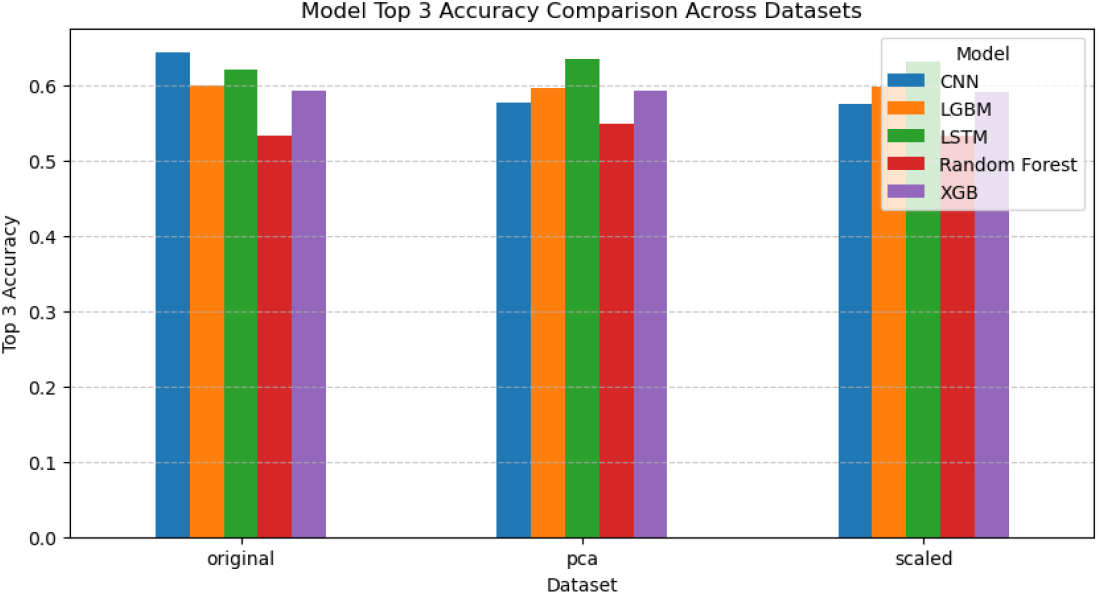
Top-3 Accuracy Comparison of Models in 3 datasets

**Fig 8.**
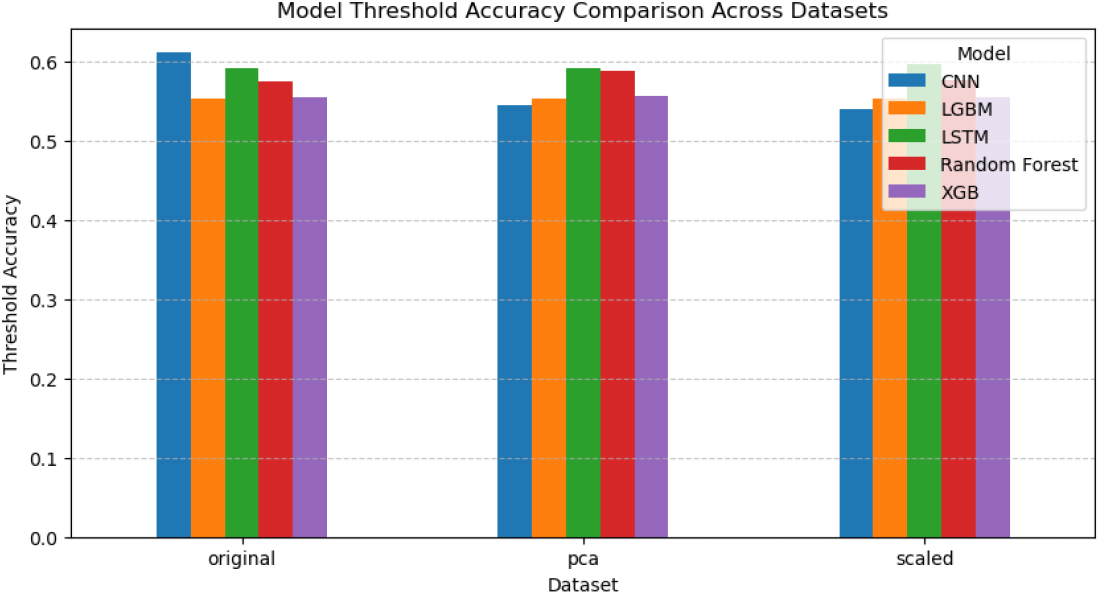
Threshold Accuracy Comparison of Models in 3 datasets

##### Accuracy Comparison

Figure 6 shows the overall accuracy of each model across the datasets. Key observations include that LSTM almost consistently achieved the highest accuracy across all datasets, indicating its ability to model sequential dependencies in the data. Random forest performed closely behind LSTM, suggesting that tree-based methods might be as effective on these datasets or under these preprocessing conditions. CNN displayed slightly lower accuracy compared to the other models.

##### Top-3 Accuracy Comparison

Figure 7 presents the Top-3 accuracy for the models. Notable findings are that LSTM maintained its advantage, achieving almost the highest Top-3 accuracy across all datasets, emphasizing its robustness in providing accurate predictions among its top three outputs. CNN also performed strongly, demonstrating its ability to rank predictions effectively. Random Forest exhibited a smaller margin between its accuracy and Top-3 accuracy, suggesting a less diverse set of high-confidence predictions compared to the other models.

##### Threshold Accuracy Comparison

Figure 8 highlights the threshold accuracy, where predictions within a ±1 range of the true label are considered correct.

Observations include that random forest and LSTM showed similar performance, with both models achieving high threshold accuracy across all datasets. This indicates their capability to make predictions close to the true labels even when exact matches are not achieved. CNN lagged slightly behind, but its performance improved noticeably under this metric compared to standard accuracy, suggesting its predictions are often near the true values.

##### General Trends Across Datasets

The PCA-applied dataset generally resulted in better performance for random forest compared to the original and scaled datasets.

This indicates that dimensionality reduction using PCA likely enhanced the quality of the features by removing noise and redundancies. The scaled dataset exhibited comparable performance to the original dataset, suggesting that scaling alone did not significantly alter the predictive performance.

### 2.12 Performance per label

#### Performance Comparison for Individual Labels

Figures 9, 10, and 11 provide a detailed comparison of the performance of Random Forest, LSTM, and CNN models across all labels using three evaluation metrics: threshold accuracy, Top-3 accuracy, and overall accuracy.

**Fig 9.**
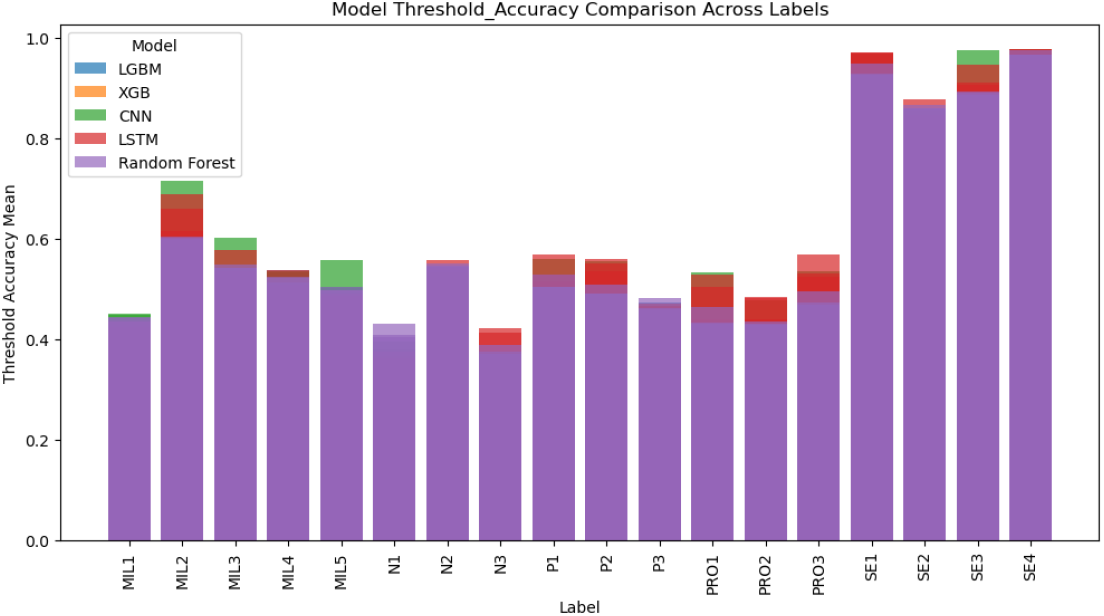
Threshold Accuracy Comparison of Models in all labels

**Fig 10.**
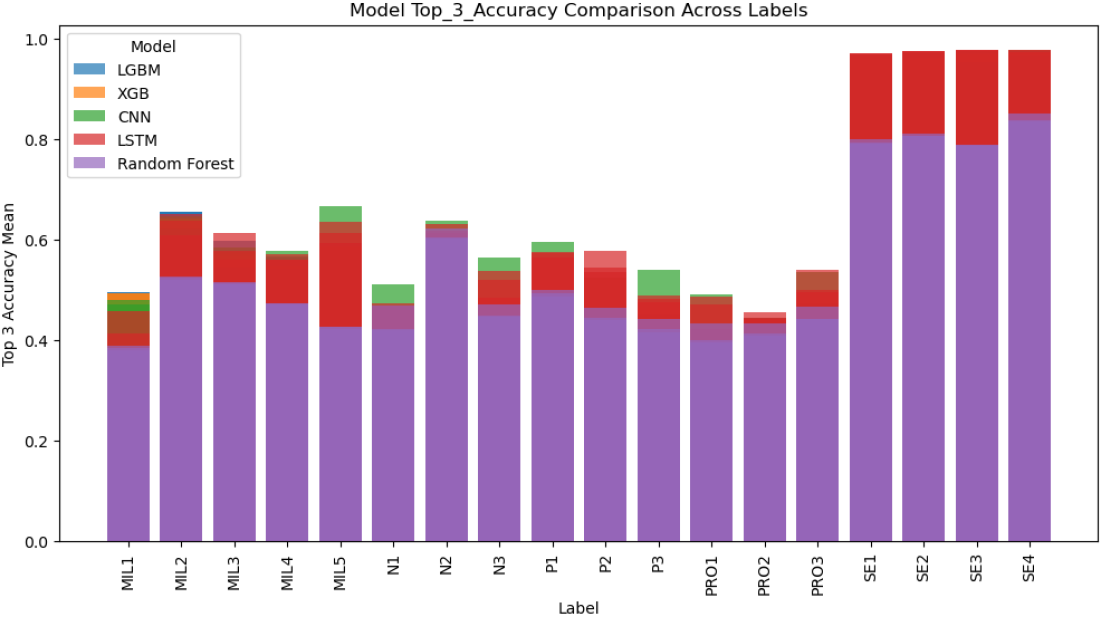
Top-3 Accuracy Comparison of Models in all labels

**Fig 11.**
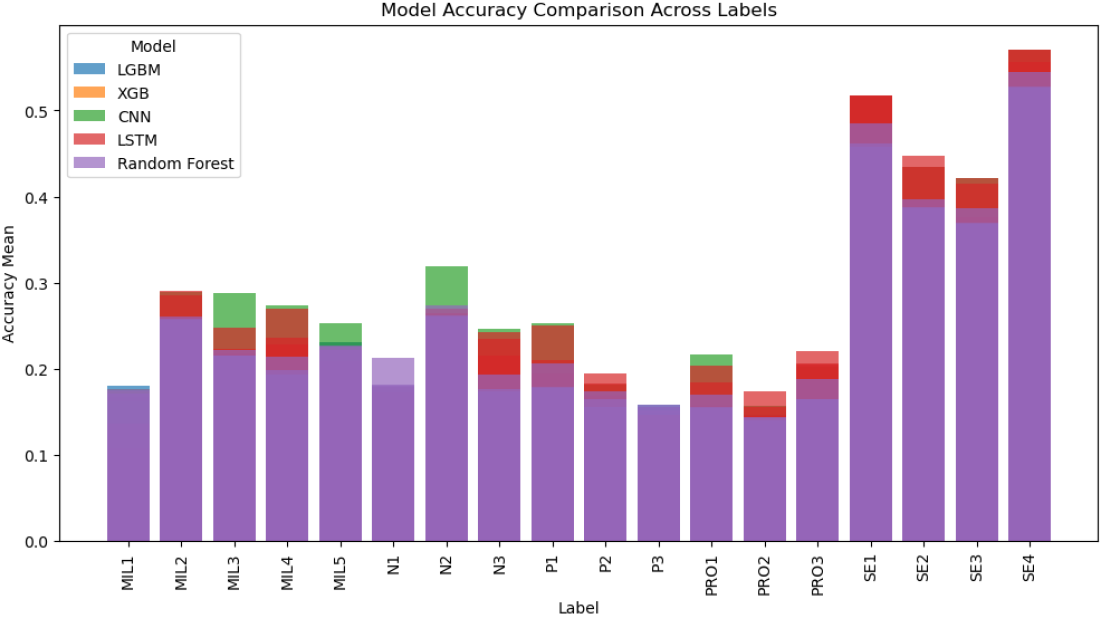
Accuracy Comparison of Models in all labels

#### Threshold Accuracy Across Labels

Figure 9 illustrates the threshold accuracy, where predictions are considered correct if they fall within a ±1 range of the true label. Key insights include that LSTM consistently achieved the highest threshold accuracy for most labels, leveraging its sequential modeling capabilities to generate accurate predictions near the true value. Random forest also performed strongly, achieving similar threshold accuracy to LSTM for several labels, particularly in categories where spatial patterns are critical. CNN exhibited slightly lower threshold accuracy across most labels, suggesting that its feature-splitting mechanism might struggle with certain label-specific complexities.

#### Top-3 Accuracy Across Labels

Figure 10 presents the Top-3 accuracy, reflecting the proportion of predictions where the true label is within the top three predictions. Observations include that LSTM achieved high Top-3 accuracy across all labels, with CNN slightly lower, indicating its robustness in ranking predictions effectively. Random Forest showed competitive Top-3 accuracy but lagged behind the other models, especially for labels associated with more nuanced patterns.

#### Overall Accuracy Across Labels

Figure 11 highlights the overall accuracy for each label. Key findings are that LSTM demonstrated the highest accuracy for almost all labels. Random Forest showed strong performance overall, with its accuracy closely matching LSTM’s for most labels. CNN displayed lower accuracy compared to LSTM and random forest but not too much, but its performance not too bad, suggesting it may excel in similar scenarios.

#### Trends Across Labels

The LSTM model performed particularly well for labels reflecting its strength in sequence modeling. The Random Forest model, while slightly less effective overall, demonstrated robust performance for certain labels. The CNN also showed comparable performances.

### 3 SUncertainty Analysis

Figure 12 presents the SHAP analysis of label prediction for MIL4 in the scaled dataset. This figure illustrates the overall feature importance across all predicted classes. The classification model organizes the response variables into multiple categories, corresponding to different psychological states. In this visualization, `acc_l2_pct_95`, `eda_tonic_rms`, `and acc_acc_x_mean` emerge as the most influential features, with consistently high SHAP values.

**Fig 12.**
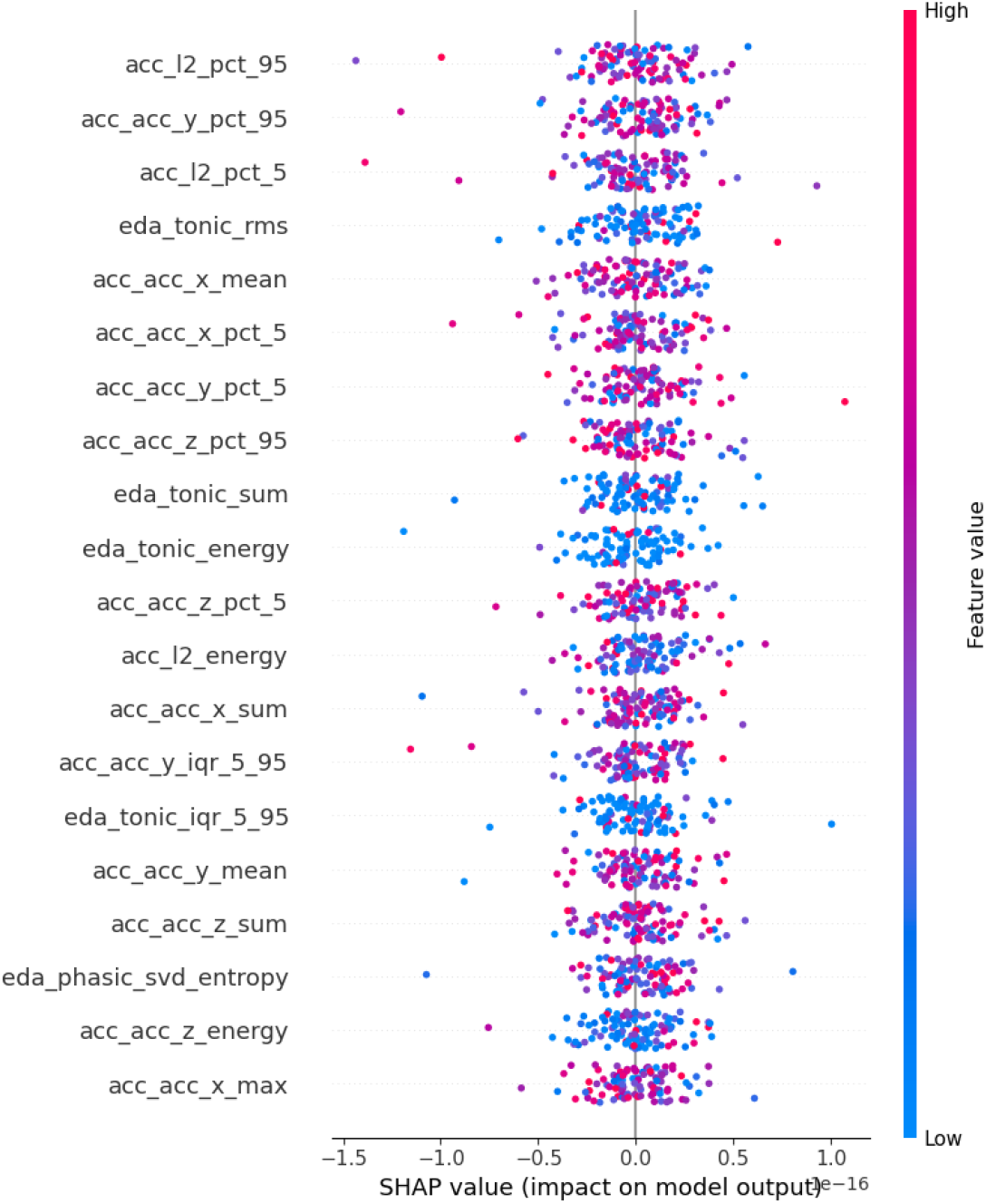
Shap analysis of MIL4 for random forest on scaled dataset. acc refers to accelerometer-based features, eda refers to electrodermal activity-based features,

Notably, eda tonic rms plays a significant role in higher-class predictions, while `acc_l2_pct_95` and `acc_acc_y_pct_95` exhibit strong contributions across the entire label range. Features such as `acc_acc_x_pct_5` and `eda_tonic_energy` exhibit moderate but consistent importance, suggesting their role as secondary predictors. This figure highlights the dominance of movement-related features (`acc_*`) in classification decisions, along with the notable impact of electrodermal activity features (`eda_*`). The results shown in this plot apply to the scaled dataset. In general, across most labels, SHAP consistently highlighted ACC features as the most influential, particularly those capturing overall movement intensity and variation. EDA features also contributed meaningfully, especially in labels linked to emotional arousal. HRV features were less dominant but occasionally appeared in top contributors for labels like Self-Esteem or Meaning in Life. These trends are reflected in the SHAP plot for MIL4, which was selected as a representative example.

To assess model uncertainty and calibration, we applied MAPIE to generate class-wise prediction intervals for five models—XGBoost, LGBM, LSTM, CNN and Random Forest—across 18 psychological-state labels on the raw dataset. The MAPIE-based interval plots revealed substantial differences in both confidence and calibration across models and labels. Tree-based models (LGBM, XGBoost, and Random Forest) generally showed clearer separation between correct (green) and incorrect (red) predictions along the probability midpoint axis, with wider intervals for uncertain samples and narrower ones for confident, correct predictions. XGBoost and Random Forest, in particular, exhibited strong calibration, with most incorrect predictions clustered at lower probability midpoints and correct predictions at higher ones. In contrast, the CNN model displayed minimal separation and lower probability midpoints overall, suggesting underconfidence and poor discriminability in its outputs. LSTM showed moderate calibration, though with slightly more mixed patterns across labels. Across labels, some (e.g., MIL1, SE2) were consistently handled well by most models, while others (e.g., N1, P3) showed systematic misclassifications, evident in large uncertainty intervals and high error rates across all models. These MAPIE results highlight both model- and label-specific strengths and weaknesses and provide actionable insights for improving robustness through targeted refinement or ensemble calibration strategies. MAPIE class-wise prediction intervals for the five models—XGBoost, LGBM, LSTM, CNN and Random Forest are shown in Figures 13, 14, 15, 16, and 17, respectively.

**Fig 13.**
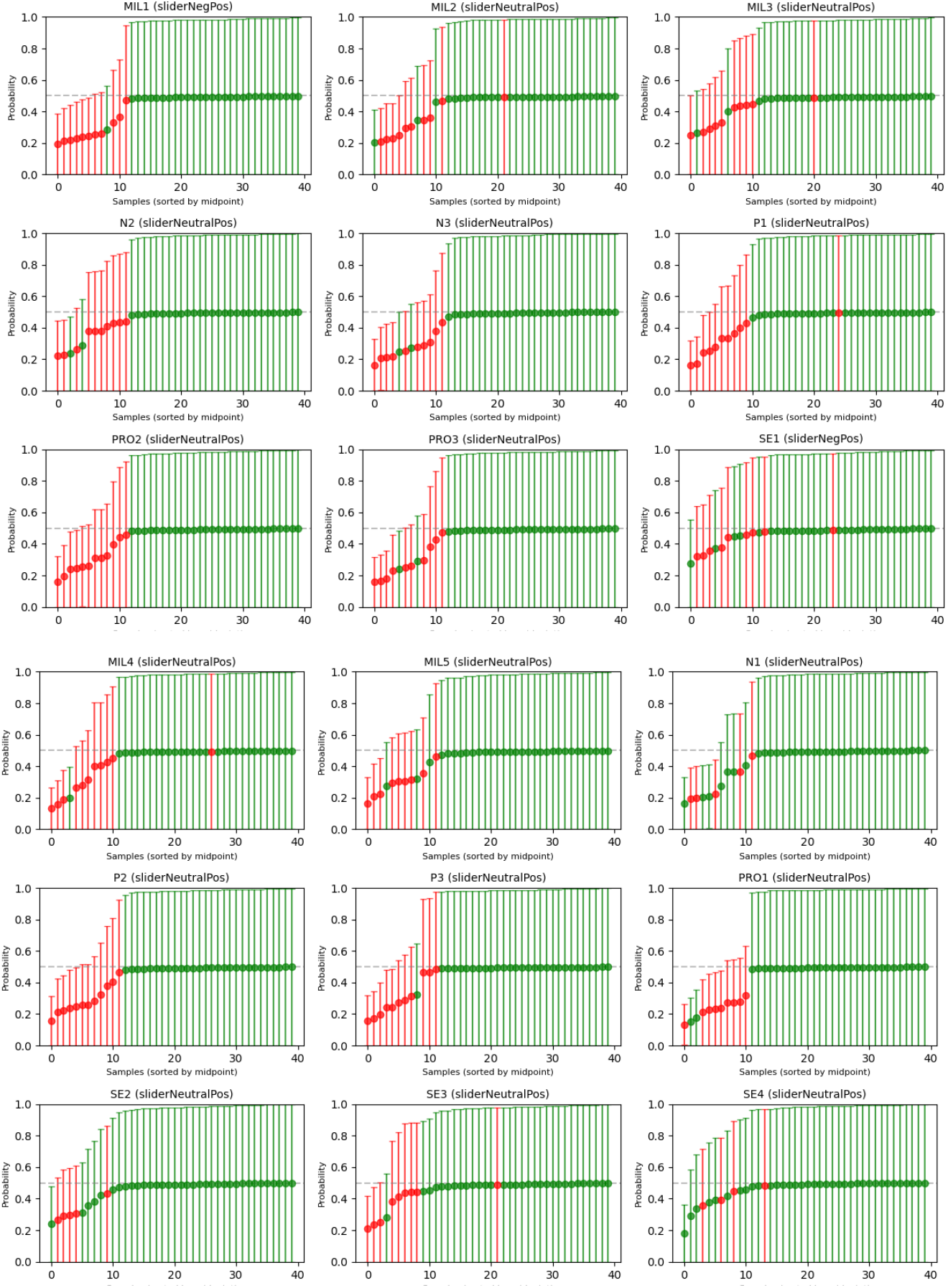
MAPIE plots for the XGBoost model for all the labels.

**Fig 14.**
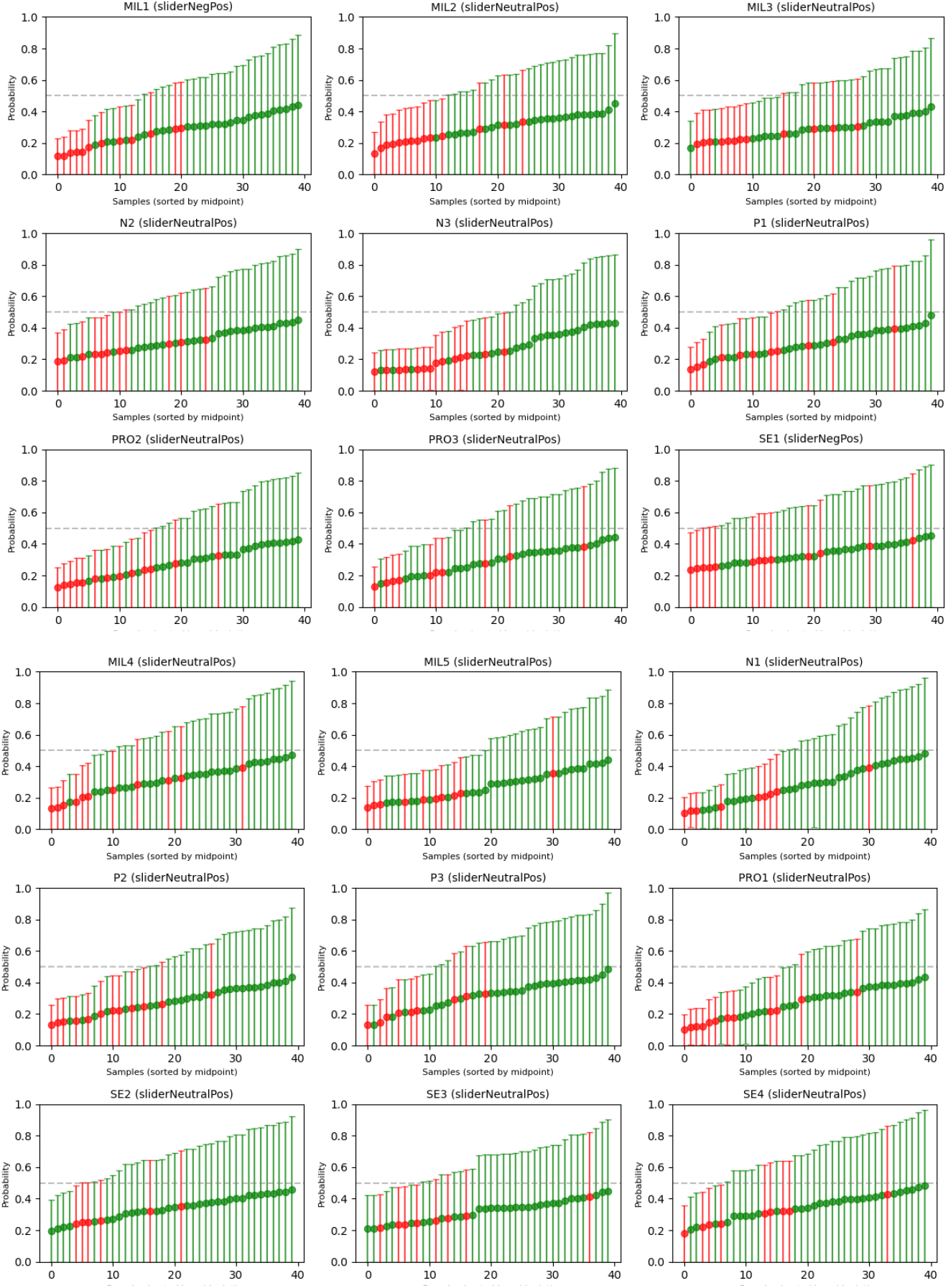
MAPIE plots for the LightGBM model for all the labels.

**Fig 15.**
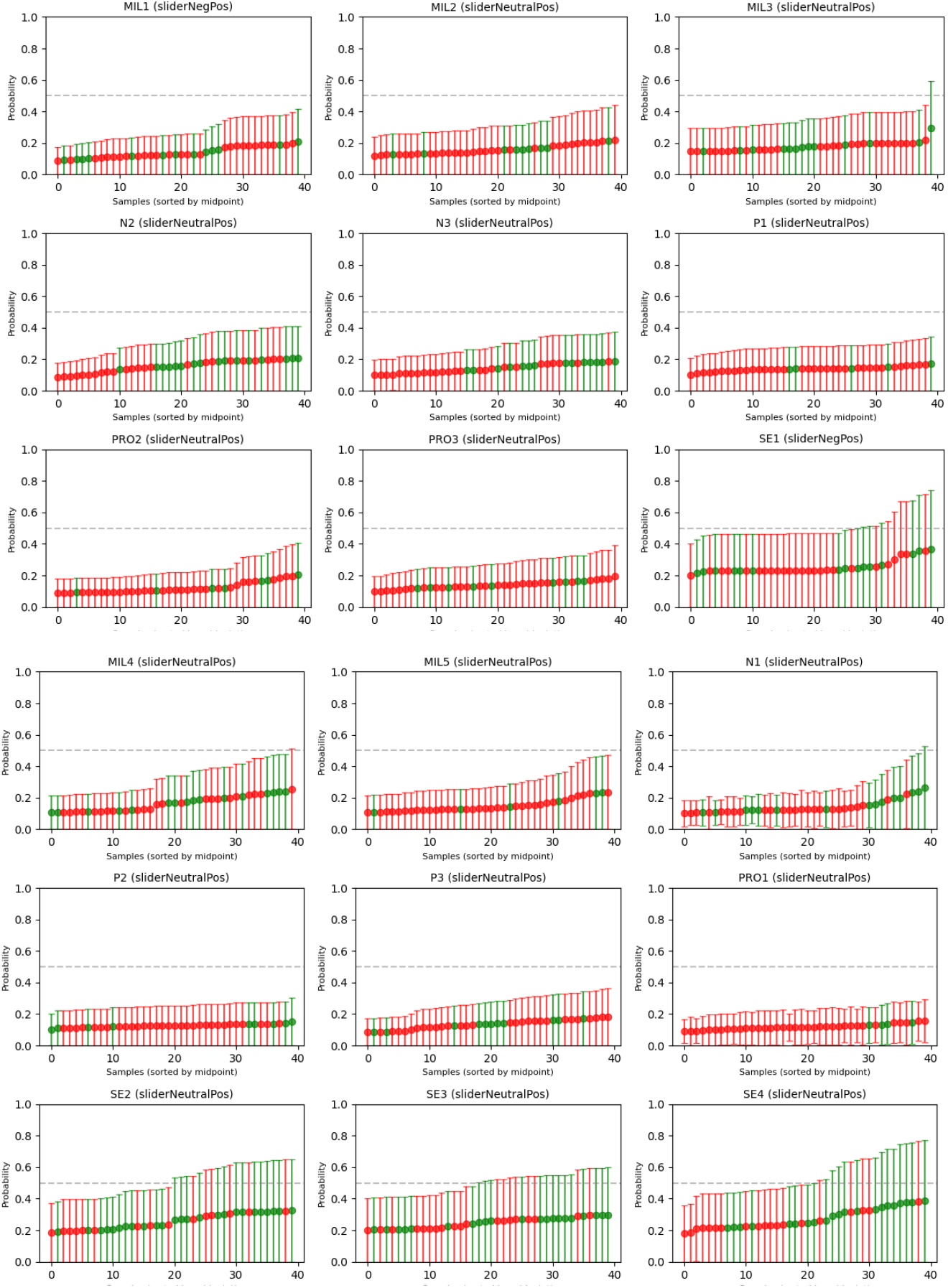
MAPIE plots for the LSTM model for all the labels.

**Fig 16.**
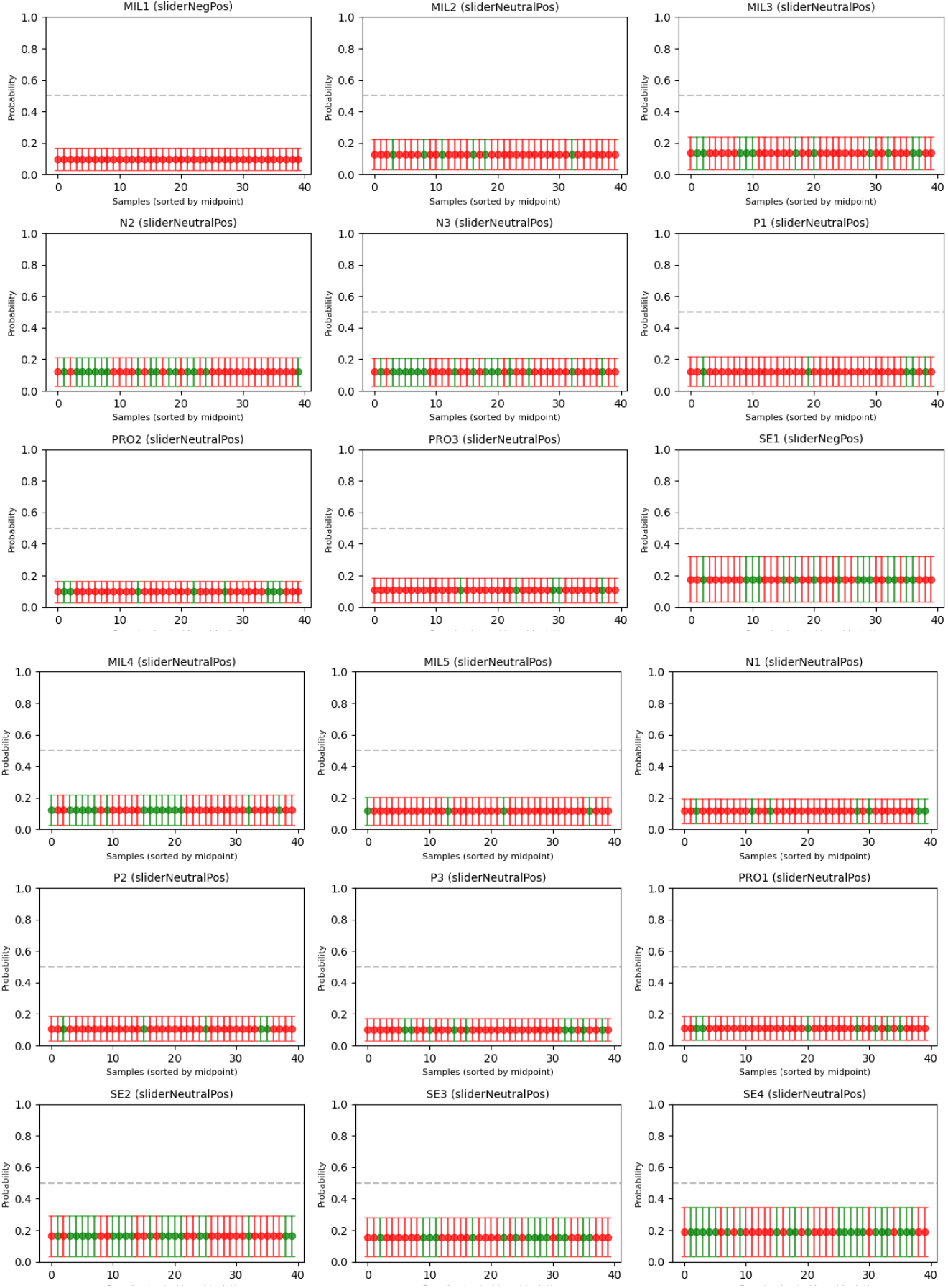
MAPIE plots for the CNN model for all the labels.

**Fig 17.**
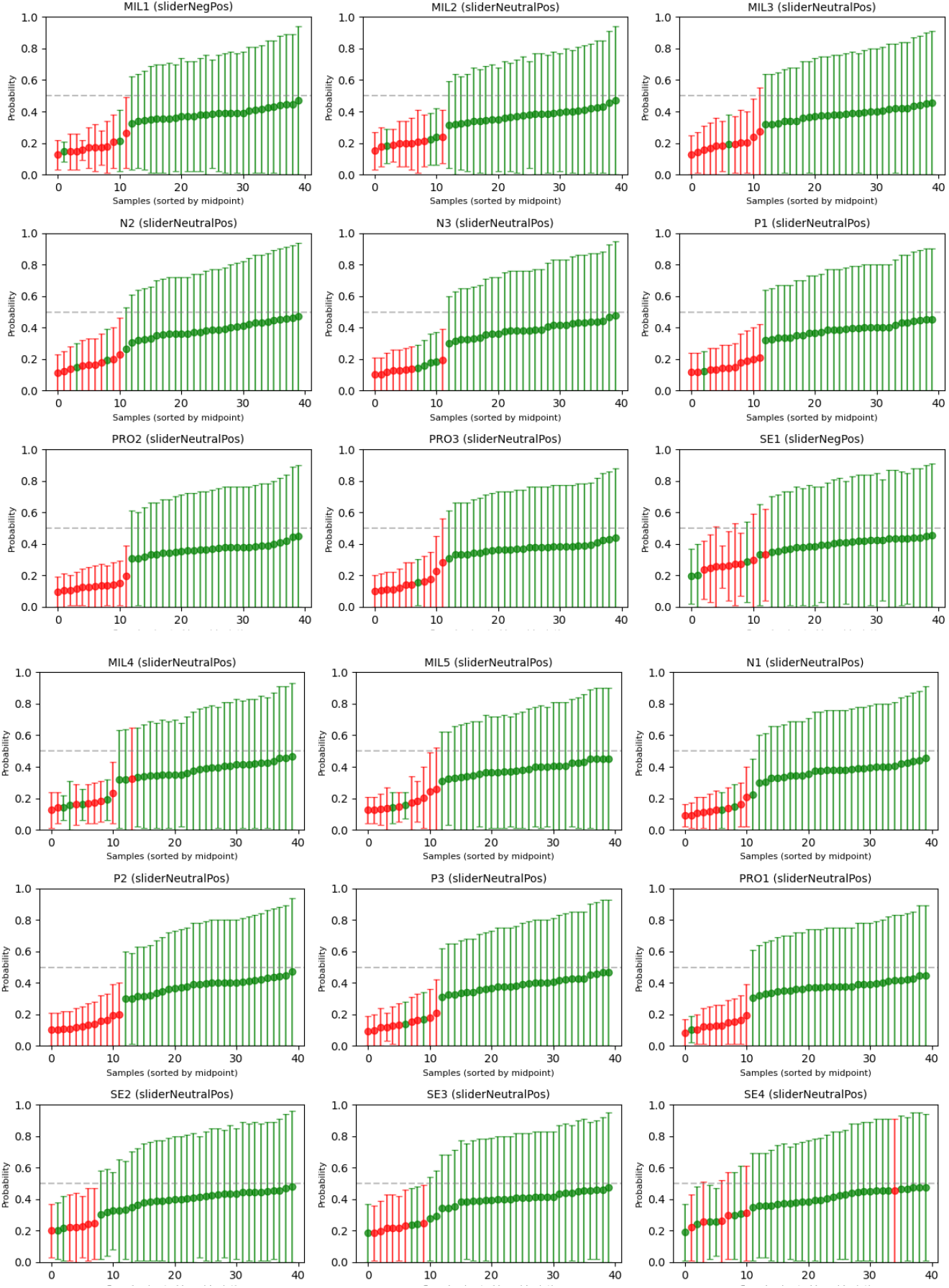
MAPIE plots for the Random Forest model for all the labels.

## Discussion

In this study, we investigated whether machine learning and digital phenotyping could accurately predict positive psychological states, such as self-esteem, personal meaning, and the quality of interpersonal relationships, measured through EMA over eight days among 34 participants. We observed marked variability in predictive performance across different constructs (placeholder for min, max, average). Using SHAP feature analysis, we found that accelerometer-derived metrics emerged consistently as the most informative features for predicting these positive states, underscoring the critical role of movement and physical activity in everyday psychological experiences.

When comparing our model’s predictive accuracies to those reported in the literature for mental illnesses such as depression, several notable differences and similarities emerge. Typically, previous studies investigating depression have employed between 31 and *>* 20000 participants (median = 108) and were conducted over periods ranging from 10 days to several months [22]. Notably, only a few studies have examined whether digital biomarkers can predict psychological states measured via EMA, whereas most research has focused on their predictive value for trait-like constructs—such as depression—assessed through questionnaires [22]. One persistent challenge highlighted in this body of research is that prediction models often experience decreased accuracy when applied to larger and more heterogeneous samples [23]. In our case, the sample predominantly consisted of young, highly educated, and physically healthy individuals, representing a relatively homogeneous cohort. Consequently, caution should be exercised in generalizing our findings beyond this specific demographic group.

Our finding highlighting the importance of movement as captured by accelerometer data aligns with a broader body of evidence suggesting a meaningful relationship between physical activity and mental health. Recent meta-analytic evidence indicates that dance-based therapy and mild cardiovascular exercise can yield outcomes comparable to or even surpassing cognitive-behavioral therapy (CBT) or pharmacological interventions for depression [24]. Extending these findings to our study, it appears plausible that physical activity not only alleviates negative emotional states but may also actively foster positive psychological states. Therefore, accelerometer-derived digital biomarkers hold significant promise for developing personalized interventions to promote physical and mental health in real-world contexts [25].

A particularly novel contribution of our study was the application of conformal prediction methods to generate prediction intervals for digital phenotyping outcomes [26]. We found (placeholder sentence summarizing main findings regarding prediction intervals, such as the average interval size, variables with largest and smallest intervals). The availability of reliable prediction intervals has considerable clinical relevance, offering clinicians and healthcare providers more transparent, interpretable predictions and informing confidence levels associated with model outputs. Moreover, prediction intervals derived from conformal prediction methods may facilitate real-time decision-making processes, enabling interventions such as JITAIs that can operate with guaranteed accuracy bounds and distribution shifts [26].

Several limitations should be acknowledged in interpreting these findings. First, despite high data compliance overall, heart rate variability (HRV) measurements were particularly sensitive to motion artifacts, a well-known limitation in physiological research that resulted in a significant number of missing values. Additionally, although EMA compliance was high, the quality of self-reported positive psychological states might be improved by employing validated EMA scales, such as adaptations of the PERMA model [27]. Future studies should aim to address these limitations by expanding the sample size and study duration, incorporating other types of sensor data (e.g., smartphone-based metrics), and recruiting more diverse populations, including clinical samples that may exhibit lower levels of positive psychological states (e.g., individuals with anhedonia or depression). Real-time monitoring and the integration of personalized interventions, such as prompting movement-based behavioral activation after detecting prolonged sedentary periods, represent promising next steps for advancing this research into applied clinical practice.

## Conclusion

In conclusion, this feasibility study demonstrates that digital phenotyping combined with machine learning can meaningfully predict positive psychological states, underscoring movement data’s particular utility in capturing emotional well-being. Our findings further illustrate the importance of incorporating transparency in model predictions through conformal prediction intervals, facilitating their integration into personalized, real-time adaptive interventions. Addressing methodological limitations and broadening the scope of future studies will be crucial for translating these technologies into reliable and effective tools for enhancing mental health and well-being in diverse populations.

## Data Availability

Data cannot be shared publicly because of its potentially sensitive nature and because no explicit consent was obtained for sharing the data.

## Notes

### Competing Interest Statement

The authors have declared no competing interest.

### Funding Statement

The author(s) received no specific funding for this work.

### Author Declarations

The Ethics Committee of the University of Twente's Faculty of Behavioural Sciences approved this study (project 240133, obtained on 23/02/2024). Written informed consent was obtained from all participants

